# SARS-CoV-2 seroassay optimization and performance in a population with high background reactivity in Mali

**DOI:** 10.1101/2021.03.08.21252784

**Authors:** Issaka Sagara, John Woodford, Alassane Dicko, Amatigue Zeguime, M’Bouye Doucoure, Jennifer Kwan, Irfan Zaidi, Justin Doritchamou, Maryonne Snow-Smith, Nada Alani, Jonathan Renn, Ivan Kosik, Jaroslav Holly, Jonathan Yewdell, Dominic Esposito, Kaitlyn Sadtler, Patrick Duffy

## Abstract

Serological tests are an indispensable tool to understand the epidemiology of the SARS-CoV-2 pandemic, particularly in areas where molecular diagnostics are limited. Poor assay performance may hinder the utility of these tests, including high rates of false-positivity previously reported in sub-Saharan Africa. From 312 Malian samples collected prior to 2020, we measured antibodies to the commonly tested SARS-CoV-2 antigens and four other betacoronaviruses by ELISA, and assessed functional cross-reactivity in a subset by SARS-CoV-2 pseudovirus neutralization assay. We then evaluated the performance of an ELISA developed in the US, using two-antigen SARS-CoV-2 spike protein and receptor-binding domain. To optimize test performance, we compared single and two-antigen approaches using existing assay cutoffs and population-specific cutoffs for Malian control samples (positive and negative). Background reactivity to SARS-CoV-2 antigens was common in pre-pandemic samples compared to US controls (43.4% (135/311) for spike protein, 22.8% (71/312) for RBD, and 33.9% (79/233) for nucleocapsid protein). SARS-CoV-2 reactivity correlated weakly with other betacoronavirus reactivity, varied between Malian communities, and increased with age. No pre-pandemic samples demonstrated functional activity. Regardless of the cutoffs applied, specificity improved using a two-antigen approach. Test performance was optimal using a two-antigen assay with population-specific cutoffs derived from ROC curve analysis [Sensitivity: 73.9% (51.6-89.8), Specificity: 99.4% (97.7-99.9)]. In the setting of high background reactivity, such as sub-Saharan Africa, SARS-CoV-2 serological assays need careful qualification is to characterize the epidemiology of disease, prevent unnecessary harm, and allocate resources for targeted control measures.

## INTRODUCTION

Multiple serological tools have been developed to monitor SARS-CoV-2 spread during the COVID-19 pandemic. These tools are important to understand the epidemiology of disease through serosurveillance, particularly in areas where gold-standard molecular diagnostics are limited. To optimize test performance, serological tools may require adaptation for use in the target population and using locally available laboratory infrastructure. These issues disproportionately affect lower income nations, where reliable data are scarce and serosurveillance may be useful to direct the limited Public Health resources and to motivate equitable global vaccine distribution.

Serological assays that measure SARS-CoV-2 antibodies typically target one of three viral antigens: spike protein, nucleocapsid protein (NCP), or the receptor binding domain (RBD), a fragment of spike protein mediating viral adhesion [2, 3]. Despite the large number of assays developed, the majority have not been qualified for use in populations where demographics and exposure histories differ significantly from the original assay validation population in high income countries. This is particularly relevant in sub-Saharan Africa, where high rates of false positivity have been described for many serological assays [4-6]. Recently, high rates of false positivity were reported using commercial SARS-CoV-2 serological assays in Benin, Tanzania and Zambia, conceivably due to cross-reactivity with other coronaviruses, or non-specific polyclonal reactivity that may be associated with unrelated conditions, including malaria [7, 8]. False positive tests may cause unnecessary alarm and harm at an individual and population level. As a result, in the setting of background reactivity, particular attention must be given to test specificity. Specificity may be improved by adjusting thresholds for seropositivity or by testing for antibodies to multiple SARS-CoV-2 antigens, as currently recommended by the US FDA, particularly in low-prevalence settings [9].

To understand and optimize the performance of serological tools for population serosurveillance in Mali, we evaluated pre-pandemic samples collected from Malian cohorts prior to 2020 for background reactivity to the commonly tested SARS-CoV-2 antigens, cross-recognition with other betacoronaviruses, and *in vitro* SARS-CoV-2 pseudovirus neutralizing activity. We subsequently evaluated the performance of an assay developed in the US using a Malian negative control (pre-pandemic) and positive control (convalescent) samples. Finally, we optimized assay performance by comparing single and dual antigen approaches and establishing Malian population-specific cutoffs.

## MATERIALS AND METHODS

### Plasma Samples

Pre-pandemic negative control plasma samples collected from Malian individuals (n=312) were obtained from ongoing studies at four MRTC sites with a strong history of community engagement in malaria research conducted in collaboration with NIAID/NIH (NCT02942277, NCT03952650, NCT03989102, NCT01322581). The four geographically distinct contributing sites were Sotuba, an urban center study population of healthy adults in urban Bamako; Bancoumana, a rural study population of healthy adults located along a major road transport route between Mali and Guinea; Ouelessebougou, a rural study population of women in their childbearing years located along a major road transport route between Mali and two neighboring countries, Burkina Faso and Cote d’Ivoire; and Kalifabougou, a rural study population of all ages. Rural centers experience high rates of seasonal malaria infection. All samples were collected prior to 2020.

Pre-pandemic US plasma samples (2017, n=20) were collected from healthy adults aged 18-50 through an existing NIH study (NCT03083847).

All pre-pandemic samples were collected during NIH-sponsored studies that were approved by the Malian USTTB FMPOS human research ethics committee and the NIAID/NIH institutional review board and were conducted in accordance with the Declaration of Helsinki and Good Clinical Practice guidelines.

Convalescent PCR-confirmed COVID-19 US positive control samples (n=10) were provided by the Adventist Hospital, Maryland. De-identified residual clinical samples for non-human subject research were obtained in accordance with 45 CFR 46.

Convalescent PCR-confirmed COVID-19 positive control samples were collected from Malian individuals (n=23) following discharge from Point G Hospital. At the time of collection, local management guidelines mandated hospital-based isolation of all confirmed cases, irrespective of severity. Convalescent samples were collected as part of a Public Health surveillance activity in collaboration with the Malian Ministry of Health COVID-19 Coordination Unit and with the approval of the USTTB FMOS-FAPH ethics committee (No2020/114/CE/FMOS/FAPH).

### Defining SARS-CoV-2 antigen background reactivity Malian samples

To identify the most suitable SARS-CoV-2 antigens for population serosurveillance in Mali, we examined pre-existing reactivity to SARS-CoV antigens and to antigens of other coronaviruses in the negative control samples from Mali. Provisional thresholds for positivity were established for each antigen from a small cohort of pre-pandemic US samples (n=20), calculated as the mean plus three standard deviations of these results.

Negative control samples were tested by ELISA at LMIV/NIAID, Bethesda, for antibodies to SARS-COV-2 spike protein, RBD and NCP, and the spike proteins of SARS-CoV-1, MERS-CoV, OC43 and HKU1 (Table 1). Data were assessed by site and by age group to understand possible demographic and geographic factors affecting antigen reactivity. Potential cross-recognition between SARS-CoV-2 and other betacoronavirus spike antigens was examined by linear correlation. Functional activity of reactive antibodies in a subset of samples was assessed by pseudovirus neutralization assay.

**Table 1:** Negative control cohort for antibody testing: population characteristics and sample size

| Site | Number of samples (N) | Date of collection | Sex, female (n) | Age group, years (n) |  |  | Antibody testing (n) |  |  |  |
| --- | --- | --- | --- | --- | --- | --- | --- | --- | --- | --- |
|  |  |  |  | <10 | 10-17 | ≥18 | SARS-CoV-2 spike | SARS-CoV-2 RBD | SARS-CoV-2 NCP | Other B-CoV spike |
| Sotuba | 64 | January 2017 | 20 | 0 | 0 | 64 | 64 | 64 | 0 | 0 |
| Bancoumana | 81 | May 2019 | 28 | 0 | 0 | 81 | 80 | 81 | 81 | 81 |
| Ouelessebougou | 67 | May 2019 | 67 | 0 | 0 | 67 | 67 | 67 | 67 | 67 |
| Kalifabougou | 100 | May 2018 | 59 | 61 | 9 | 29 | 100 | 100 | 85 | 100 |

Due to limitations in sample volume, some samples were not able to undergo all antibody testing.
RBD: receptor binding domain, NCP: nucleocapsid protein, Other B-CoV: other betacoronaviruses

### SARS-CoV-2 antigens and other betacoronavirus spike proteins

SARS-CoV-2 full-length spike protein (VRC-SARS-CoV-2 S-2P-3C-His8-Strep2×2 [10]) and RBD protein (Ragon-SARS-CoV-2 S-RBD(319-529)-3C-His8-SBP [11]) and the full-length spike proteins for the other betacoronaviruses SARS-CoV-1, MERS-CoV, OC43 and HKU1 [12] were produced as previously described.

Full length SARS-CoV-2 nucleocapsid protein was prepared at LMIV/NIAID, Bethesda. Nucleocapsid protein (GenBank: QHD43423.2, amino acids 1-419) was synthesized and cloned (GenScript) into the AgeI and KpnI sites in the expression vector pHLSEC [13]. Endotoxin free plasmid used for nucleocapsid protein production was generated using a high-speed maxi prep kit (Qiagen) and the protein was produced in the Expi293 cell line (Thermo Fisher). Typically, Expi293 suspension cells in Expi293 Expression medium (Thermo Fisher) were grown at 37°C and 8% CO2, maintaining cultures at continuous log phase growth (3.0-5×10^6^) for 3-4 passages after thawing. The day before transfection, 500 mL of culture was seeded at a density of 2.5-3×10^6^ cells/mL in a 2 L flask. The day of transfection, cells were diluted back to 2.5-3×10^6^ prior to transfection. Expi293 cells were transfected using 1.4 mL of ExpiFectamine (Thermo Fisher) and 0.5 mg of plasmid DNA per 0.5 L of cells. Plasmid DNA was diluted into 25 mL of OptiMEM (Thermo Fisher) and filter sterilized through a 0.2 micron filter. The ExpiFectamine was slowly added to 25 mL of OptiMEM, gently mixed and incubated for 5 min. The diluted ExpiFectamine was then added slowly to the diluted DNA, gently mixed and incubated at RT for 10-20 min. The mixture was added to the cells slowly while swirling the flask. The flask was returned to the incubator at 37°C and 8% CO2 for 16-20 hrs. The following day, both enhancer I and II (Thermo Fisher) were added to the Expi293 cultures and returned to the incubator.

Cultures at 4 days post-transfection were centrifuged at 10,000 x g for 30 min. The spent culture media was sequentially filtered through 0.45 µm and 0.2 µm filters. The clarified spent media was loaded on a 5 mL HisTrap Excel NTA column (GE Life Sciences). The column was washed with 20 column volumes of wash buffer (20 mM sodium phosphate, 0.5 M NaCl, 0 to 30 mM imidazole, pH 7.4). The bound protein was eluted with a step gradient of elution buffer (20 mM sodium phosphate, 0.5 M NaCl, 500 mM imidazole, pH 7.4). The eluted NTA fractions were confirmed to contain nucleocapsid protein by SDS-PAGE (4-12% Bis-tris, Thermo Fisher) and Coomassie staining. The fractions were pooled, concentrated, and buffer exchanged by diafiltration using a 10 kDa cutoff centrifugal filter unit (Millipore Sigma). The absorbance at 280 nm was determined and the concentration of the recombinant nucleocapsid protein was calculated from the predicted extinction coefficient from the protein sequence. Proteins were then aliquoted and stored at −80°C.

### Enzyme-Linked Immunosorbent Assay

As MRTC has significant expertise performing ELISAs and local infrastructure supports this platform, the semi-automated two-antigen ELISA developed at NIBIB/NIH [14] was selected for transfer from the US to Mali. Preliminary process qualification was conducted at LMIV/NIAID, Bethesda prior to transfer and validation of the assay at the MRTC/DEAP Immunology Laboratory in Bamako, Mali.

ELISAs were performed as previously described [12, 14]. In brief, 100 µL of antigen suspension in 1xPBS was added to each well of a 96-well Immulon 4 HBX ELISA plate and allowed to coat overnight at 4°C for 16 hours. Antigen suspension concentrations were SARS-CoV-2 spike protein 1 µg/mL; RBD 2 µg/mL; NCP 1 µg/mL; and other betacoronavirus spike proteins 1 µg/mL. Wells were washed three times with 300 µL of 1xPBS + 0.05% Tween20 by automatic plate washer followed by blocking for 2 hours at room temperature with 200 µL of 1x PBS + 0.05% Tween20 + 5% nonfat skim milk. After blocking, wells were washed again three times. Heat inactivated plasma samples were diluted at 1:400 in blocking buffer and 100 µL of sample added to wells. Positive, negative, and blank controls were included on all plates. Positive controls were monoclonal antibody dilutions of the neutralizing antibody CR3022 (10 µL of 500, 250, 125, 62.5, 31.3 ng/mL, LMIV internal construct). Negative controls were pooled pre-pandemic plasma from Malian children diluted and added to wells as for samples. All samples and controls were run in duplicate. Samples were incubated for 1 hour at room temperature, then washed three times with 300 µL of wash buffer. One hundred (100) microliters of goat anti-Human IgG (H+L) Cross-Adsorbed secondary antibody, HRP (ThermooFisher) diluted at 1:4000 in blocking buffer was added to wells, incubated for 1 hour at room temperature, then washed three times with 300 µL of wash buffer. One hundred (100) microliters of 1-Step Ultra TMB Substrate (ThermoFisher) was added and the plate was incubated for 10 minutes prior to the addition of 1N sulfuric acid stop solution (ThermoFisher). Absorbance was read at 450 nm and 650 nm on the Spectramax M3 microplate reader (Molecular Devices, US). Absorbance values (optical density, OD) were collected at 450 and 650 nm. A650 was subtracted from A450 to remove background signal.

### Pseudovirus neutralization assay

SARS-CoV-2 spike glycoprotein (Sgp) neutralization was measured by pseudotyped VSVdG-EGFP-SARS2-Sgp flow cytometry neutralization assay at LVD/NIAID, Bethesda. Forty microliter aliquots of sample dilutions in 40 µL of DMEM (Gibco) supplemented with 8% FBS (Hyclone), 10 mM HEPES (Corning), and 50 µg/mL gentamicin (Sigma) (diluent solution) was added to each well of a non-tissue culture treated polypropylene 96 round-bottom well plate (Corning). Serial sample dilutions were prepared in half-log10 increments starting at 1:20. Pseudotyped VSVdG-EGFP-SARS2-Sgp virus in diluent solution (40 µL at MOI=0.05) was then added to each well. Positive control sample dilutions from de-identified US convalescent PCR-confirmed COVID-19 cases kindly provided by the Adventist Hospital, Maryland, and recombinant neutralizing antibody H4 a-RBD hIgG1 [15]) were prepared in a similar fashion. The pseudotyped virus-sample mixture was incubated at 37°C for one hour in 9% CO2. Forty thousand BHK21-ACE2 cells dissociated with 0.5% Trypsin EDTA, no-phenol red (Gibco), and diluted in DPBS (Gibco) were transferred to each well of a new non-tissue culture treated polypropylene 96 round-bottom well plate (Corning). The DPBS was gently aspirated after one round of centrifugation at 2,000 RPM for five minutes. Seventy microliters of pseudovirus-sample mixture was immediately added to wells containing BHK21-ACE2 cells and incubated overnight at 37°C in 9% CO2. Following incubation, cells were sedimented by centrifugation at 2,000 RPM for five minutes. The pseudovirus-sample mixture was gently aspirated and 10 µL of 0.5% Trypsin EDTA, no-phenol red (Gibco) was added and incubated with cells for two minutes at room temperature. Cells were resuspended in 100 µL HBSS (Gibco) supplemented with 0.1% BSA. Samples were analyzed using a BD Celesta instrument employing a high throughput system unit. We performed the analysis performed using FlowJo software (TreeStar). The frequencies of EGFP-positive cells were normalized by the non-Ab–treated samples and fitted to nonlinear regression curves using the dose-response inhibition model for neutralization assay with GraphPad Prism7 software.

### BHK-21-ACE2 cell production

The BHK-21 cell line was maintained in DMEM (Gibco, 10566016) supplemented with 10% FBS, grown at 37°C, 9% CO2 humid atmosphere and tested mycoplasma-negative using Universal Mycoplasma Detection Kit (ATCC 30-1012K). Human ACE2 cDNA cloned to pCDNA3.1(+) was kindly provided by Sonja Best (NIAID/NIH). The open reading frame of ACE2 was amplified in PCR using Q5 Hot Start High-Fidelity 2X Master Mix (New England Biolabs) using primers flanked with SfiI restriction enzyme recognition sequence (underlined) followed by irrelevant nucleotides to facilitate digestion of PCR product. Primers: hACE2_Fw: 5’-T<u>GGCCTGACAGGCC</u>CTAAAAGGAGGTCTGAACATCATC-3’, hACE2_Rv: 5’-T<u>GGCCTCTGAGGCC</u>ACCATGTCAAGCTCTTCCTGGC-3’. PCR product was purified using DNA Clean & Concentrator-5 (Zymo Research), sequentially digested with FastDigest SfiI and FastDigest DpnI (Thermo Scientific) and the cloning insert purified by DNA Clean & Concentrator-5 (Zymo Research). Transposon vector pSBbi-BH, kindly provided by Eric Kowarz (Addgene plasmid #60515), was chosen for generation of a cell line stably expressing ACE2 taking advantage of Sleeping Beauty transposase system. Plasmid pSBbi-BH was digested by FastDigest SfiI (Thermo Fisher Scientific), dephosphorylated using FastAP Thermosensitive Alkaline Phosphatase (1 U/µL) (Thermo Scientific), size-selected by 1% agarose electrophoresis and purified using Zymoclean Gel DNA Recovery Kit (Zymo Research). Prepared insert and vector fragments were ligated using Rapid DNA Ligation Kit (Roche), transformed to One Shot TOP10 Chemically Competent *E. coli* (Invitrogen) and selected on carbenicillin LB plates. The sequence of the selected clone was confirmed by Sanger sequencing. Transposase vector pCMV(CAT)T7-SB100 was kindly provided by Zsuzsanna Izsvak (Addgene plasmid #34879), designated further as SB100X. BHK-21 cells were seeded at 5 x 10^5^ cells/ml density in a 60 mm tissue-culture-treated dish in a total volume of 4 mL complete media 24 hours prior transfection. Plasmids pSBbi-BH hACE2 and SB100X were co-transfected in a molar ratio of 10:1 using TransIT-LT1 Transfection Reagent (Mirus Bio) according to manufacturer instructions. Twenty-four hours post transfection the cells were split into a T-75 TC flask and selected with 250 μg/mL hygromycin (Invivogen) for two weeks. The surface expression of ACE2 was confirmed by flow cytometry using anti-human ACE2 AlexaFluor 647-conjugated antibody (R&D Systems, FAB9332R) (Supplementary Figure 1). The expression was further confirmed by western blot using ACE2 Antibody (Cell Signaling Technology, #4355).

**Supplementary Figure 1:**
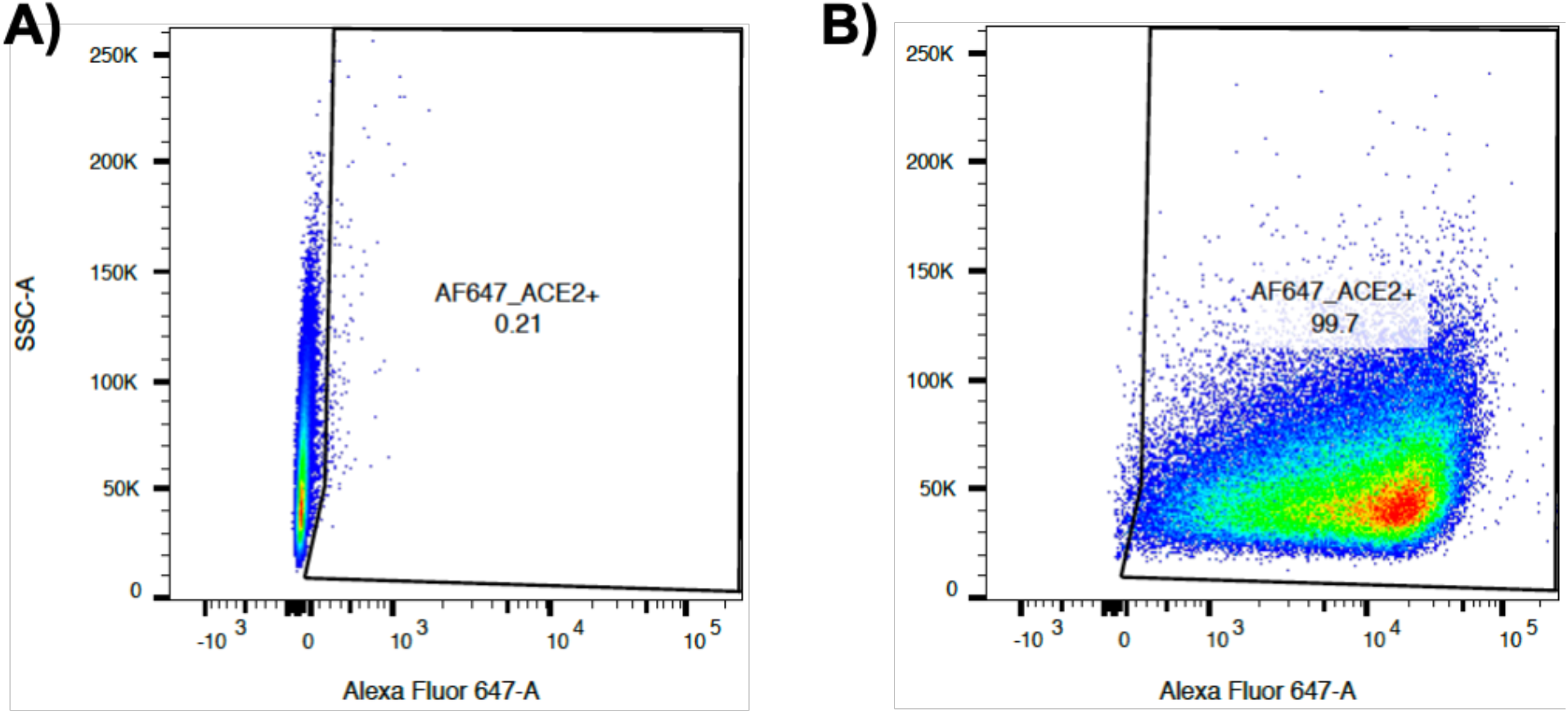
Confirmation of BHK-21 cell ACE2 expression by flow cytometry using anti-hACE2 AlexaFluor 647-conjugated antibody. A) BHK-21 control B) BHK-21 BH-hACE2

### VSVdG-EGFP-SARS2-Sgp virus production

To generate non-replicative self-reporting VSVdG-EGFP-SARS2-Sgp, we employed a commercial VSVdG system (Kerafast). Briefly, a T75 tissue flask was seeded with five to seven million 293T cells (60-70% confluency) in DMEM (Gibco) supplemented with 8% FBS (Hyclone) and 10mM HEPES (Corning). The next day 50 µL FuGene low-cytotoxicity transfection reagent (Promega) and 2 mL OPTIMEM media (Gibco) were combined (FuGene mixture) and 32 µg of pHEF-VSV-Gp plasmid DNA (VRC/NIAID) and 2 mL OPTIMEM media (Gibco) (DNA OPTIMEM mixture) were combined. Both mixtures were incubated at room temperature for five minutes. The FuGene and pDNA OPTIMEM mixtures were then combined and incubated at room temperature for 20 −30 minutes (transfection mixture). Media from the seeded T75 flask was gently aspirated and replaced with 14 mL of the fresh DMEM (Gibco) supplemented with 8% FBS (Hyclone) and 10 mM HEPES (Corning). The transfection mixture was then added dropwise and incubated with the cell culture for 48 hours at 37°C in a CO2 incubator. Virus mixture was prepared by combining 0.3 mL of VSVdG-EGFP-Gp P2 (titer ∼10e6/mL) and 1.7 mL of DMEM (Gibco) supplemented with 8% FBS (Hyclone) and 10 mM HEPES (Corning). Two milliters of medium was aspirated from the T75 flask cell culture containing transfection mixture and gently replaced with virus mixture and incubated at 37°C for one hour in a CO2 incubator while gently rocking every 10-15 minutes. Following incubation, media was aspirated and 13 mL of DMEM (Gibco) supplemented with 8% FBS (Hyclone) and 10 mM HEPES (Corning) was added. After 36-40 hours of incubation at 37°C in a CO2 incubator, harvested media was centrifuged at 4,700 RPM for 5 minutes. VSVdG-EGFP-Gp P3 were snap-frozen in 1ml aliquots in an isopropanol-dry ice mixture and stored at −80°C. VSVdG-EGFP-VSV-Gp P3 pseudotyped virus was titrated on BHK21 cells. Serial two-fold dilutions of VSVdG-EGFP-VSV-Gp pseudotyped virus in 0.1 mL DMEM (Gibco) supplemented with 8% FBS (Hyclone) and 10mM HEPES (Corning) were added to 100,000 BHK21 cells. After incubating cells overnight, the frequency of EGFP positive cells was measured by flow cytometry using a BD Celesta instrument. We calculated titer using the formula: infectious titer/ml=frequency of EGFP positive cells x cell number (100,000) x dilution of viral stock x inverse value of inoculum volume (0.1 mL). The average of three data points from an unsaturated fraction of the titration curve was calculated as the titer.

To generate VSVdG-EGFP-SARS2-Sgp, a T75 tissue flask was seeded with eight million BHK21-SARS2-Sgp-CTd16aa cells in DMEM (Gibco) supplemented with 8% FBS (Hyclone) and 10 mM HEPES (Corning). VSVdG-EGFP-Gp P3 aliquots were thawed on ice and diluted in 2 mL of DMEM (Gibco) supplemented with 8% FBS (Hyclone) and 10mM HEPES (Corning) to MOI=4. Media was aspirated from the T75 flask, and the virus inoculum transferred to BHK21-SARS2-Sgp-CTd16aa cells before incubating in a CO2 incubator for 75 minutes, gently rocking the flask every 15 minutes. Following incubation, the virus inoculum was carefully aspirated and replaced with 10 mL of pre-warmed DMEM (Gibco) supplemented with 8% FBS (Hyclone) and 10 mM HEPES (Corning) before incubating the flask in a CO2 incubator at 37°C. Cells were inspected by fluorescence microscopy and once most of the cells turned green-fluorescent and rounded (30-36 hours), media was harvested. The suspension was centrifuged at 4,700 RPM for 10 minutes at 4°C and the supernatant removed. Pelleted cells were resuspended in 1-2 ml media, freeze-thawed three times, and sonicated on ice at 50% amplitude five times in one-second pulses. The mixture was then centrifuged at 4,700 RPM for 10 minutes at 4d°C. Supernatant, combined with media supernatant, was collected and aliquoted and snap-frozen in isopropanol-dry ice mixture for 20 minutes before storing at −80°C. VSVdG-EGFP-SARS2-Sgp pseudotyped was titrated on BHK21-ACE2 cells by flow cytometry as for VSVdG-EGFP-VSV-Gp pseudotyped virus. To exclude the contribution of potentially recycled VSV-Gp, we compared the susceptibility to infection of wtBHK21 and BHK21-ACE2 cells and infectious titer in the presence and absence of the 1E9F11 and 8G5F11 anti-VSV-Gp antibody mixture (both 10 µg/mL). We concluded that the effect of potentially recycled VSV-Gg is negligible for the VSVdG-EGFP-VSV-SARS2-Sgp virus stock.

### Assessing performance of US cutoffs in Malian samples

The NIBIB ELISA-based serology assay was transferred for use at the MRTC/DEAP Immunology Laboratory, Bamako. This is a two-antigen assay to detect antibodies to SARS-CoV-2 spike protein and RBD, developed for use in US population serosurveillance studies [14]. A positive test requires reactivity to both antigens above cutoffs developed in the US population. In the US population, the estimated sensitivity and specificity are 100% (95% CI: 92.9 to 100) and 100% (95% CI: 98.8 to 100) respectively.

To assess the performance of US cutoffs in the Malian population, we calculated the test characteristics using Malian negative (n=311) and positive control samples (n=23). The sensitivity, specificity, and positive predictive value across a range of plausible population seroprevalences of the US cutoffs was calculated.

### Optimizing assay performance for use in Mali

To evaluate different methods to improve assay performance, single- and two-antigen approaches were assessed using population-specific cutoffs. Population-specific cutoffs were developed using two methods. Firstly, arithmetic thresholds of two, three, and four standard deviations above the mean of the negative control cohort were calculated for each antigen. This approach was used in the original development of the assay for use in the US population [14]. Secondly, thresholds for spike and RBD were developed by interrogating ROC curves generated from the positive and negative control cohorts. Test performance characteristics of these population-specific cutoffs were calculated as for the US cutoffs.

### Data analysis

Data were analyzed using Microsoft Excel and GraphPad Prism7 software. In pre-pandemic Malian samples, the coefficient of variation for SARS-CoV-2 antigen assay absorbance values was calculated to assess variability in background reactivity in this population. Assay absorbance values for SARS-CoV-2 antigens and other betacoronavirus spike proteins were compared between demographic groups using Kruskal-Wallis tests. No adjustments have been made for multiple comparisons. Linear correlation between assay absorbance values for viral antigens and assay absorbance and neutralizing activity was assessed by Pearson correlation, as previously described [12]. Correlations were classed as negligible r<0.3, weak 0.3<r<0.5, moderate 0.5<r<0.7, or strong >0.7 [16]. Provisional test cutoffs were derived from the mean plus two, three, or four standard deviations of results from the negative control cohort, and manually interrogating the inflection points of ROC curves generated using the results of the negative and positive control cohorts. Measures of test performance are presented with 95% confidence intervals.

## RESULTS

### Pre-pandemic reactivity to SARS-CoV-2 and other coronaviruses in Mali

Negative control samples collected from four study sites between 2017 and 2019 were tested for antibodies to SARS-COV-2 spike protein (n=312), RBD (n=311) and NCP (n=233) and the spike proteins of SARS-CoV-1, MERS-CoV, OC43 and HKU1 (n=248) (Table 1). Some sample volumes were insufficient to test all antigens.

Many Malian samples demonstrated marked reactivity to the SARS-CoV-2 antigens commonly used in clinical serological tests relative to provisional thresholds derived from a small cohort of pre-pandemic US samples (n=20) (Figure 1). Background reactivity in Malian samples exceeded the mean plus three standard deviations of the pre-pandemic US sample group in 43.4% (135/311) for spike protein, 22.8% (71/312) for RBD, and 33.9% (79/233) for NCP. In Malian samples, the assay absorbance signal coefficient of variation for spike protein was 86.2%, RBD 148.2% and NCP 109.2%. This degree of pre-existing reactivity was thought likely to affect the effectiveness of serosurveillance. As a result, the characteristics of samples with high background reactivity were examined further.

**Figure 1:**
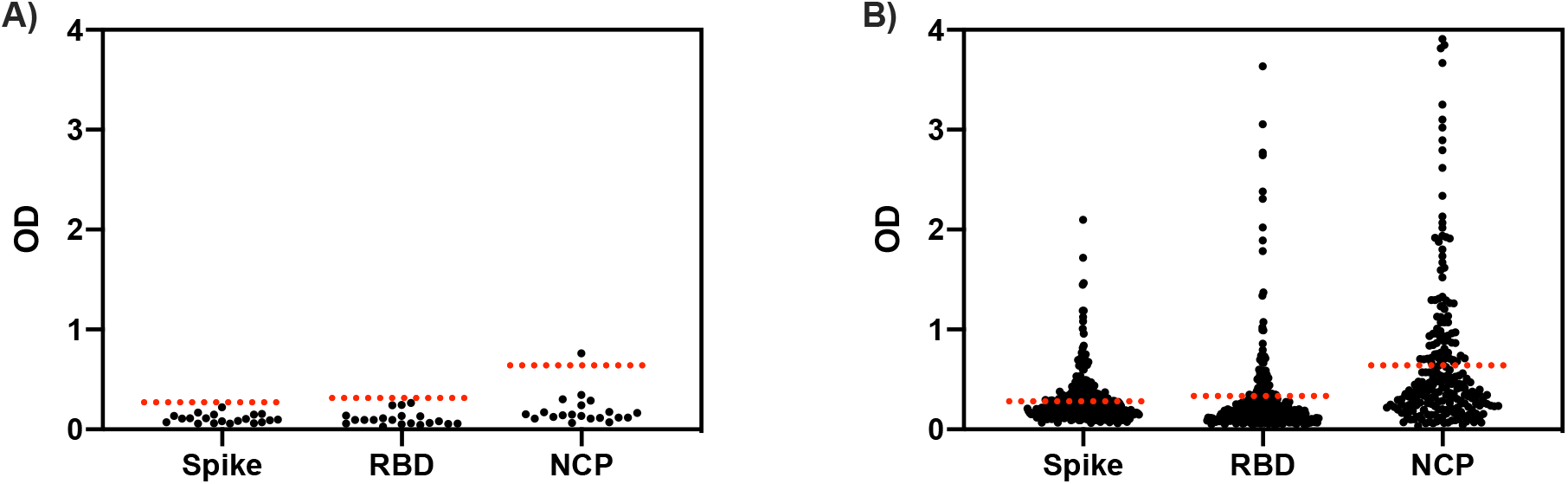
SARS-CoV-2 antigen reactivity by ELISA in COVID-19 naïve samples. A) US adult samples (n=20) B) Malian samples: Spike protein (n=311), RBD (n=312), NCP (n=233). OD: optical density, RBD: receptor binding domain, NCP: nucleocapsid protein Dotted lines represent provisional thresholds of mean plus three standard deviations of US pre-pandemic samples in panel A. US adult samples: Coefficient of variation: spike protein 40.9%, RBD 63.5%, NCP 79.7%. Malian samples: Coefficient of variation: spike protein 86.2%, RBD 148.2%, NCP 109.2%.

Pre-existing reactivity to SARS-CoV-2 spike protein, RBD and NCP antigens varied according to the site of collection (Kruskal-Wallis test, p<0.0001 for spike protein and RBD, p=0.0238 for NCP; Figure 2A-C). Reactivity was highest in samples from women in their reproductive years at the Ouelessebougou site. Samples from the Ouelessebougou site were collected at the same time of year as samples from the Bancoumana and Kalifabougou sites, reducing the potential impact of seasonal variation (Table 1). In samples from the Kalifabougou site that included children and adults (n=100), reactivity to all SARS-CoV-2 antigens increased with age group (Kruskal-Wallis test, p<0.0001 for spike protein, RBD, and NCP; Figure 2E-G).

**Figure 2:**
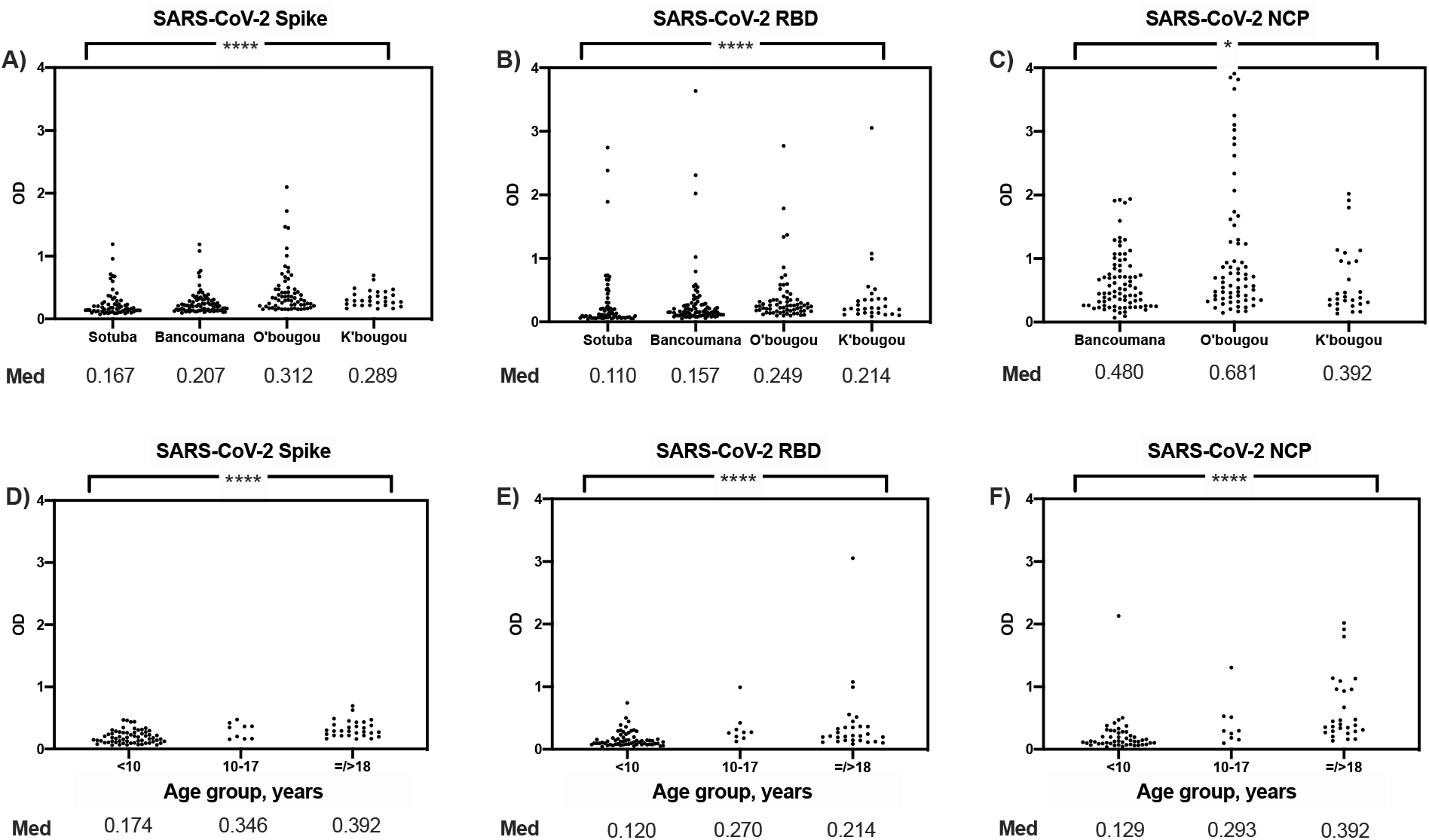
SARS-CoV-2 antigen reactivity by site (age=/>18 years) in COVID-19 naïve Malian samples. A) Spike protein B) RBD C) NCP. SARS-CoV-2 antigen reactivity by age group (Kalifabougou site) D) Spike protein E) RBD F) NCP. Med: median OD value, OD: optical density, RBD: receptor binding domain, NCP: nucleocapsid protein, O’bougou: Ouelessebougou site, K’bougou: Kalifabougou site **** represents p<0.0001, * represents p=0.01-0.05 using Kruskal-Wallis test.

Similar to the SARS-CoV-2 antigen background reactivity pattern, reactivity to other betacoronavirus spike proteins (n=233) differed between sites and was highest in samples from the Ouelessebougou site (Kruskal-Wallis test, p<0.0001 for SARS-CoV-1, MERS-CoV, OC43, HKU1, Supplementary Figure 2). In samples from the Kalifabougou site, reactivity to the common-cold coronaviruses OC43 and HKU1 increased with age group (Kruskal-Wallis test, p<0.0001 for each). Age-related reactivity was also observed for SARS-CoV-1 and MERS-CoV but was less pronounced (Kruskal-Wallis test, SARS-CoV-1 p=0.0725, MERS-CoV p=0.0017).

**Supplementary Figure 2:**
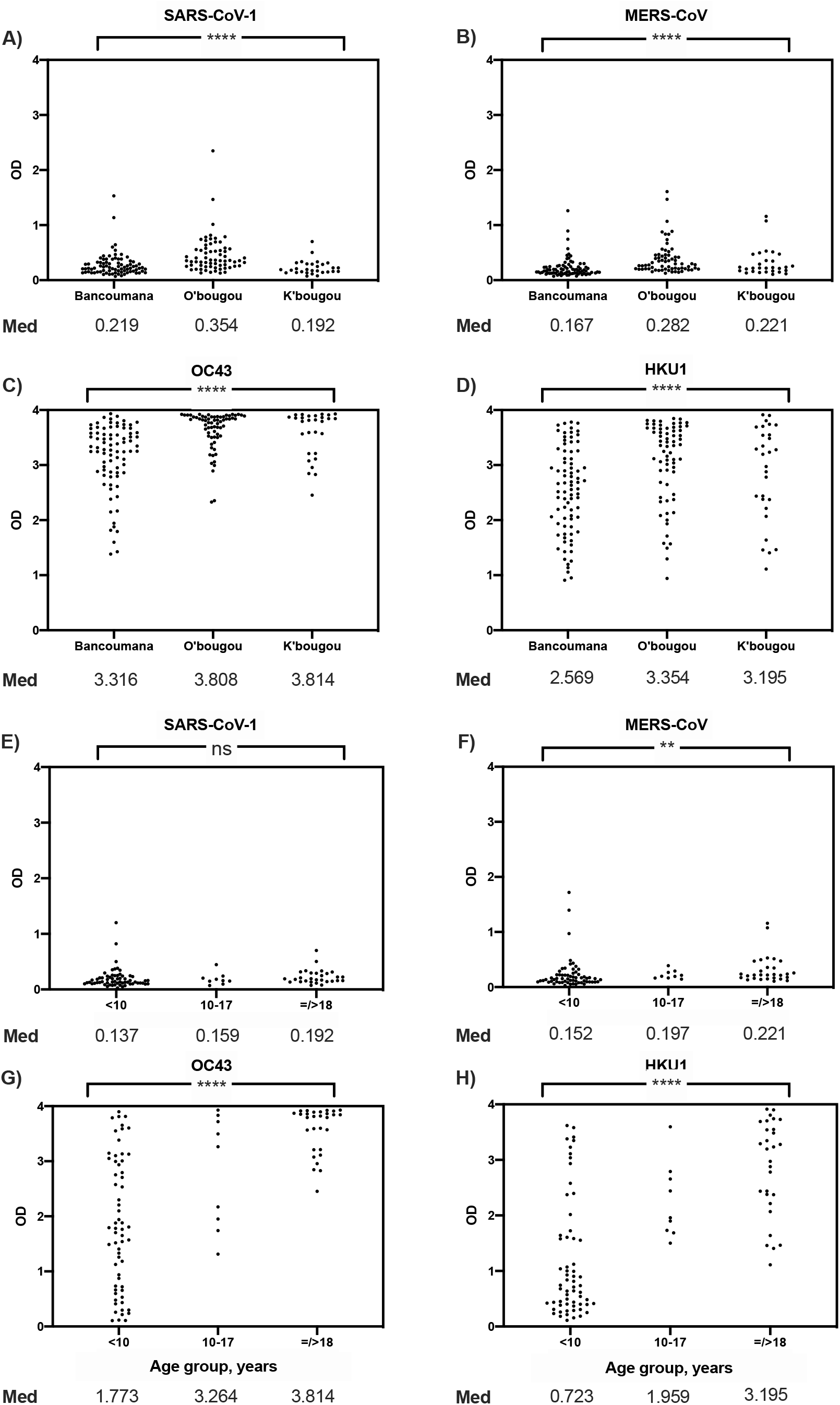
Other betacoronavirus spike protein reactivity by site (age=/>18 years) in COVID-19 naïve Malian samples. A) SARS-CoV-1 B) MERS-CoV C) OC43 D) HKU1. SARS-CoV-2 antigen reactivity by age group (Kalifabougou site) E) SARS-CoV-1 F) MERS-CoV G) OC43 H) HKU1. Med: median OD value, OD: optical density, RBD: receptor binding domain, NCP: nucleocapsid protein, O’bougou: Ouelessebougou site, K’bougou: Kalifabougou site **** represents p<0.0001, ** p=0.001-0.01, ns represents p>0.05 using Kruskal-Wallis test.

Although the patterns of site and age-related reactivity were similar for SARS-CoV-2 antigens and other betacoronavirus spike proteins, linear correlations in assay absorbance values were modest (Figure 3). In contrast, a strong correlation was observed between the common cold betacoronaviruses OC43 and HKU1 (Pearson r=0.728, p<0.0001) which share a high degree of sequence homology and may elicit serological cross-reactivity [12, 17]. SARS-CoV-2 spike and RBD reactivity correlated minimally (Pearson r=0.217, p=0.0006), despite RBD being a subunit of the whole spike protein.

**Figure 3:**
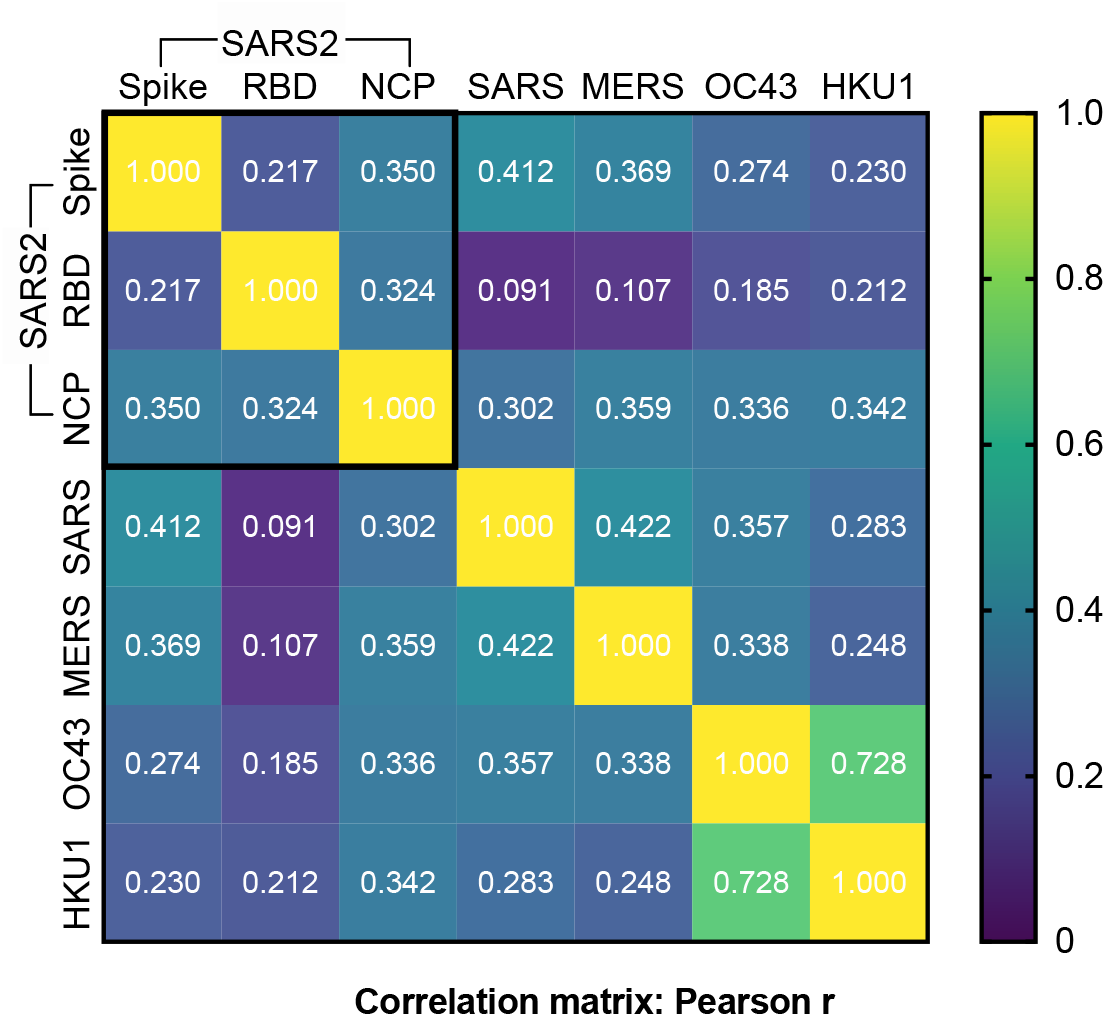
Correlation matrix of SARS-CoV-2 antigens and four other betacoronavirus spike protein reactivity in COVID-19 naïve Malian samples. SARS2: SARS-CoV-2, RBD: receptor binding domain, NCP: nucleocapsid protein, SARS: SARS-CoV-1.

To determine if SARS-CoV-2 spike protein or RBD reactive antibodies detected by ELISA in Malian pre-pandemic samples provided any functional activity, a subset of samples were tested by SARS-CoV-2 pseudovirus neutralization assay (n=89; Sotuba n=59, Bancoumana n=14, Ouelessebougou n=16). No functional activity was observed at the lowest dilution for any of the Malian samples, including samples with high assay absorbance signal to RBD, the major target of the antibody neutralization response (Figure 4). In contrast, positive control US convalescent samples (9/10) demonstrated neutralizing activity comparable to neutralizing potency of recombinant a-RBD monoconal antibody (Figure 4). In US convalescent serum samples, neutralizing activity (log10IC50) was strongly correlated with spike protein and RBD OD value (Spike protein: Pearson r=0.895, p=0.0011; RBD: Pearson r=0.841, p=0.0045).

**Figure 4:**
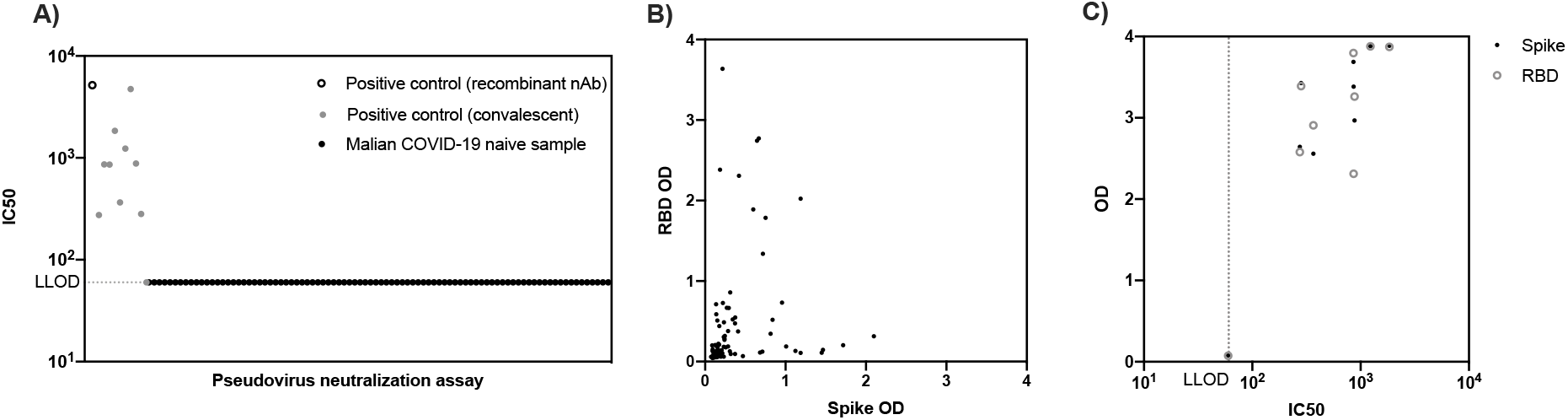
Functional activity of antibodies in COVID-19 naïve Malian samples (n=89) and positive control samples (n=11; recombinant nAb n=1, US convalescent serum n=10) A) Pseudovirus neutralization test of samples at lowest dilution (1:20) B) SARS-CoV-2 spike protein and RBD reactivity by ELISA in COVID-19 naïve Malian samples tested by pseudovirus neutralization test C) Pseudovirus neutralization test IC50 versus spike protein and RBD OD in positive control US clinical samples. IC50: half maximal inhibitory concentration, LLOD: lower limit of detection, OD: optical density, RBD: receptor binding domain, recombinant nAb: recombinant neutralizing antibody H4 a-RBD hIgG1.

### Assessing performance of US cutoffs in Malian samples

To establish the clinical effect of high background reactivity, we evaluated the performance of the NIBIB two-antigen ELISA in Malian negative control samples (n=311, four geographically distinct sites, all ages) and positive control samples (n=23, Bamako, adults) (Table 1, Supplementary Table 1). Using the assay approach and cutoffs developed in the US population, test sensitivity was 78.3% (95% CI: 56.3-92.5) and specificity was 97.4% (95% CI: 95.0-98.9) in Malian samples (Figure 5).

**Figure 5:**
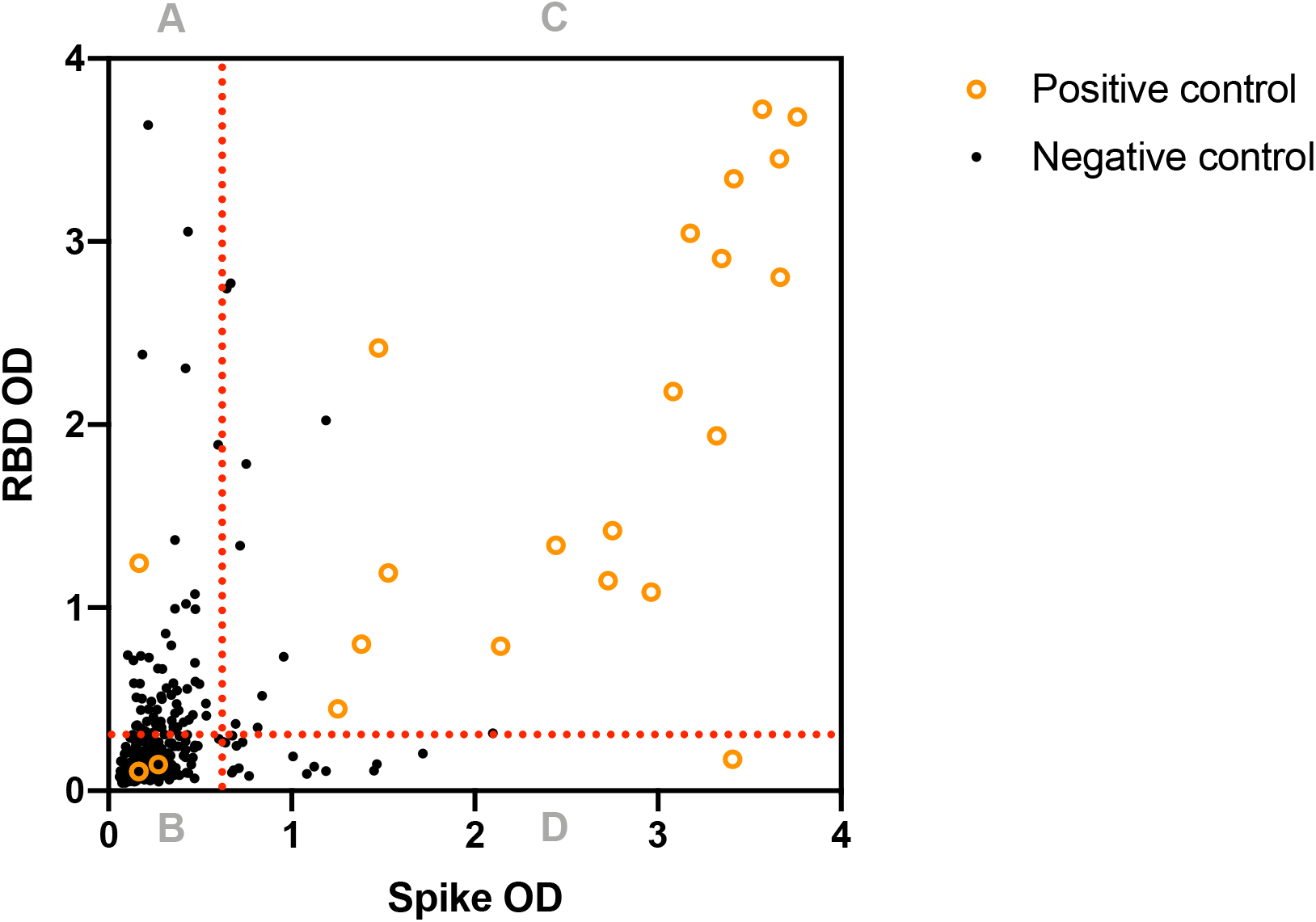
The performance of US cutoffs in Malian positive control (n=23) and negative control (n=311) samples. OD: optical density, RBD: receptor binding domain Dotted lines represent US assay cutoffs for spike protein: 0.674 and RBD: 0.306 [14]. Quadrant A (spike negative, RBD positive): positive control: 1/23, negative control: 65/311 Quadrant B (spike negative, RBD negative): positive control: 3/23, negative control: 225/311 Quadrant C (spike positive, RBD positive): positive control: 18/23, negative control: 8/311 Quadrant D (spike positive, RBD negative): positive control: 1/23, negative control: 13/311

In the negative control cohort, 8/311 (2.6%) were considered seropositive (assay absorbance signal above threshold for both spike protein and RBD). Single antigen reactivity to spike protein was observed in 21/311 (6.8%) and RBD in 73/312 (23.4%). A two-antigen approach reduced the false positivity rate several-fold compared to using any single antigen and existing US cutoffs.

In the positive control cohort, 18/23 (78.3%) were seropositive for both spike protein and RBD. This represented a modest reduction in sensitivity compared to a single antigen approach, where reactivity to spike protein alone was observed in 19/23 (82.6%) and RBD alone in 19/23 (82.6%). All subjects were adults, and the time between diagnosis by PCR and sample collection for ELISA ranged between 27 to 270 days (Supplementary Table 1). Almost half of subjects (10/23) were asymptomatic or paucisymptomatic (reporting a single symptom). Disease severity ranged from asymptomatic to critical using WHO stratification criteria [18]. Lower reactivities to spike and RBD were associated moreso with milder disease severity, rather than increased time since diagnosis (Supplementary Figure 3).

**Supplementary Table 1:** Positive control cohort population characteristics

|  |  |
| --- | --- |
| **Sex [male] (% , n)** | 74% [17/23] |
| **Age, years (median [range])** | 35 [7-74] |
| **Time since diagnosis, days (median [range])** | 181 [27-270] |
| **Severity (% , n)** |  |
| Asymptomatic | 17% [4/23] |
| Mild (no oxygen) | 57% [13/23] |
| Mod-severe (supplemental oxygen) | 9% [2/23] |
| Critical (ventilator) | 13% [3/23] |
| **Symptoms (% , n)** |  |
| Asymptomatic | 17% [4/23] |
| Paucisymptomatic | 26% [6/23] |
| Symptomatic | 57% [13/23] |

Symptoms categorized as asymptomatic, paucisymptomatic (one symptom), or symptomatic (two or more symptoms.)

**Supplementary Figure 3:**
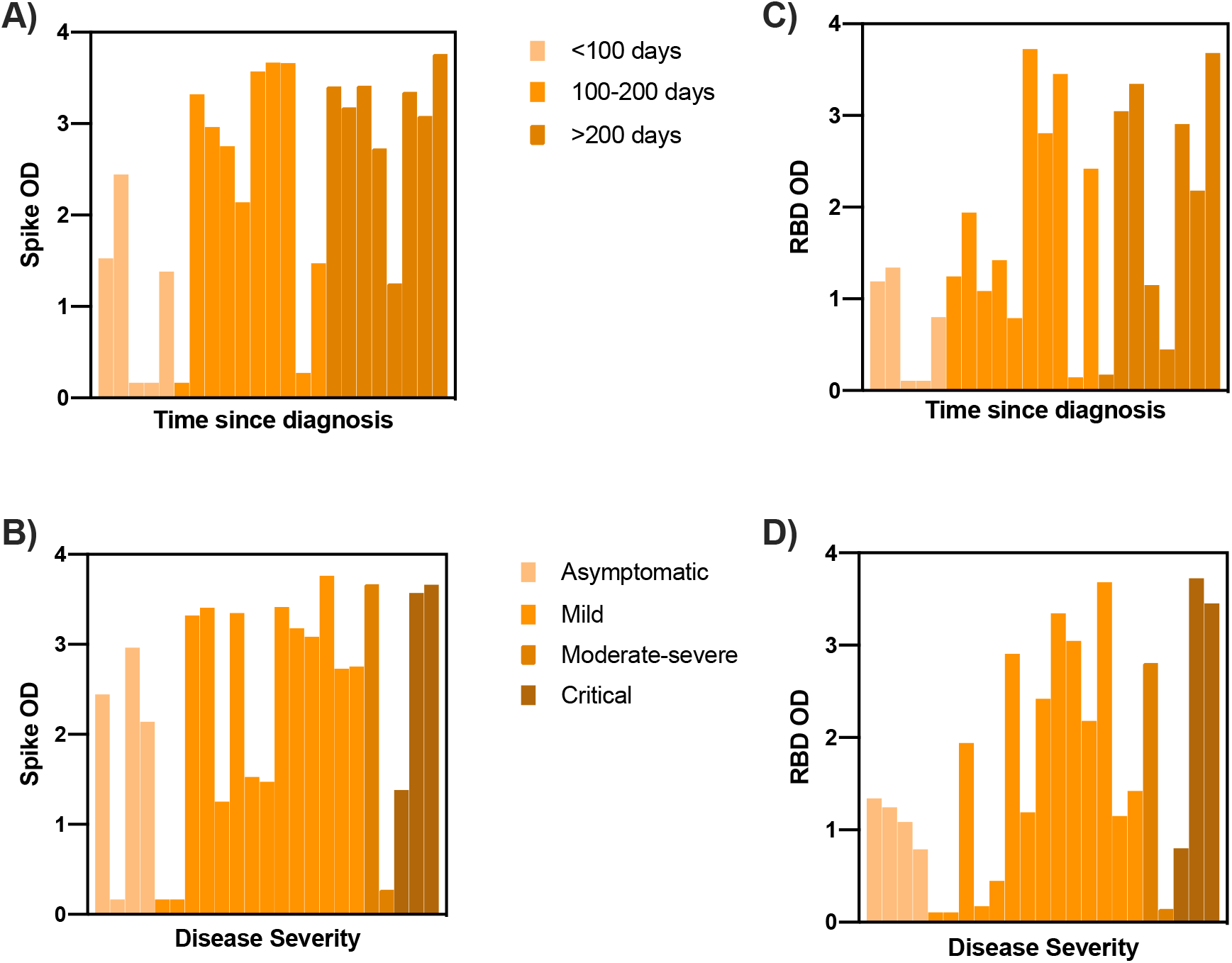
Positive control cohort population reactivity to SARS-CoV-2 spike protein by ELISA according to: A) time since diagnosis and B) disease severity. Reactivity to RBD by ELISA according to: C) time since diagnosis and D) disease severity. OD: optical density, RBD: receptor binding domain Disease severity categorized using WHO stratification criteria (Clinical Management of COVID-19 WHO, May 27 [18])

The positive predictive value of the existing assay was estimated across a range of plausible community seroprevalences. At a lower hypothetical seroprevalence of 1% the positive predictive value (PPV) was 23.5% (95% CI: 13.0-38.6) at 5% the PPV was 61.6% (95% CI: 43.9-76.6) and at 10% the PPV was 77.2% (62.3-87.4). As a result, the existing assay approach and cutoffs were not considered suitable to confidently estimate the burden of exposure in a community with limited molecular diagnostic data and an uncertain pre-test probability.

### Optimizing assay performance for use in Mali

We sought to improve test performance to better suit the Malian population. To optimize the assay, the performance of single- and two-antigen approaches was assessed using population-specific cutoffs. Population-specific cutoffs were developed using two methods. Firstly, arithmetic thresholds of two, three, and four standard deviations above the mean of the negative control cohort were calculated for each antigen. Secondly, thresholds for spike and RBD were developed by interrogating ROC curves generated for each antigen from the positive and negative control cohorts. The performance characteristics of these population-specific cutoffs were estimated in the existing cohorts (Table 2). As expected, arithmetic cutoffs improved assay specificity however this was at the expense of sensitivity. In contrast, ROC curve analysis cutoffs delivered a sensitivity of 73.9% (95% CI: 51.6-89.8) and specificity of 99.4% (95% CI: 97.7-99.9), representing an improvement on the assay cutoffs developed in a US population with a minimal loss in sensitivity. At a lower hypothetical seroprevalence of 1% the PPV was 53.7% (22.2-82.5), at 5% the PPV was 85.8% (59.8-96.1) and at 10% the PPV was 92.7% (75.9-98.1).

**Table 2:** Performance characteristics of single and two-antigen approaches using US and population-specific cutoffs in Malian positive control and negative control samples.

|  | US cutoffs | Arithmetic Malian cutoffs (mean + 2SD) | Arithmetic Malian cutoffs (mean + 3SD) | Arithmetic Malian cutoffs (mean + 4SD) | ROC curve Malian cutoffs |
| --- | --- | --- | --- | --- | --- |
| **Single antigen: Spike** |  |  |  |  |  |
| Threshold (OD) | **0.674** | **0.791** | **1.041** | **1.291** | **0.743** |
| Sensitivity (95% CI) | 82.6% (61.2-95.1) | 82.6% (61.2-95.1) | 82.6% (61.2-95.1) | 78.3% (56.3-92.5) | 82.6% (61.2-95.1) |
| Specificity (95% CI) | 92.9% (89.5-95.5) | 96.1% (93.4-98.0) | 97.4% (95.0-98.9) | 98.7% (96.7-99.7) | 95.5% (92.6-97.5) |
| **Single antigen: RBD** |  |  |  |  |  |
| Threshold (OD) | **0.306** | **1.183** | **1.625** | **2.067** | **0.766** |
| Sensitivity (95% CI) | 82.6% (61.2-95.1) | 60.9% (38.5-80.3) | 43.5% (23.2-65.5) | 39.1% (19.7-61.5) | 78.3% (56.3-92.5) |
| Specificity (95% CI) | 76.5% (71.4-81.1) | 96.5% (93.8-98.2) | 97.1% (94.6-98.7) | 98.1% (95.9-99.3) | 94.5% (91.4-96.8) |
| **Two-antigen: Spike and RBD** |  |  |  |  |  |
| Sensitivity (%; 95% CI) | 78.3% (56.3-92.5) | 56.5% (34.5-76.8) | 43.5% (23.2-65.5) | 39.1% (19.7-61.5) | 73.9% (51.6-89.8) |
| Specificity (%; 95% CI) | 97.4% (95.0-98.9) | 99.7% (98.2-100) | 99.7% (98.2-100) | 100% (98.8-100) | 99.4% (97.7-99.9) |

## DISCUSSION

If serological surveillance is to be a useful tool to aid the Public Health response to the COVID-19 pandemic, test selection and validation in the target population will be critical. Our findings highlight the need for care in SARS-CoV-2 assay selection and interpretation, and the challenge of assay implementation in sub-Saharan Africa.

Background reactivity to SARS-CoV-2 antigens was common. This reactivity varies geographically and may be in part related to prior coronavirus exposure; however other cumulative exposures and non-specific binding may also exert an effect. In the negative control cohort, reactivity to the common cold betacoronaviruses OC43 and HKU1 was common. Although MERS-CoV is thought to circulate in camel populations in Northern Mali [19] reactivity to MERS-CoV spike protein was similar to that observed for SARS-CoV-1 and SARS-CoV-2, suggesting that there is limited MERS-CoV exposure in our population. As a result, in our cohort, exposure to common cold betacoronaviruses is more likely to explain any betacoronavirus-related SARS-CoV-2 antigen cross-reactivity, rather than exposure to MERS-CoV or SARS-CoV-1. Nonetheless, we are unable to exclude the contribution of other coronaviruses, including undescribed animal coronaviruses in Mali. While cross-recognition between SARS-CoV-1 and MERS-CoV may result in SARS-CoV-2 cross-reactivity, common cold coronaviruses and SARS-CoV-2 spike are thought to have minimal cross-reactivity [12, 17].

Given the modest correlation with other betacoronavirus reactivity, it is likely that other cumulative exposures and non-specific binding contribute to SARS-CoV-2 antigen background reactivity. While understanding the cause of non-specific reactivity is less important than quantifying this reactivity, the effect of seasonal malaria warrants future assessment as it is particularly relevant in Mali. This study did not examine changes over time in the same community or assess the effect of acute malaria on SARS-CoV-2 antigen reactivity.

Although pre-existing immunity has been hypothesized as a potential explanation for the comparatively benign course of the COVID-19 pandemic in parts of sub-Saharan Africa [20, 21], we did not identify functional antibodies in Malian samples with reactivity to SARS-CoV-2 antigens. While differences in environmental exposure history and in immune response to viral challenge compared to European and North American populations may contribute to differences in diagnostic test performance and pandemic epidemiology in sub-Saharan Africa, pre-existing circulating functional antibodies were not apparent in Malian samples.

Due to high background reactivity in Malian samples, testing of a single SARS-CoV-2 antigen is unlikely to offer sufficient specificity for use in serosurveillance unless very high thresholds for positivity are applied. To overcome this high background reactivity, a two-antigen approach is more appropriate. In Mali, SARS-CoV-2 spike protein and RBD appear to be superior targets compared to NCP for such an approach. This was due to the potential for correlation with neutralizing activity [22, 23], the higher background reactivity absorbance values observed in NCP ELISA of COVID-19 naïve Malian samples, and the lower specificity of NCP as an antigen reported in other populations [24-26].

Optimizing specificity was considered the primary objective of assay adaptation, however the tradeoff with sensitivity required consideration due to the pronounced overlap between positive and negative control cohorts. The relatively low seropositivity irrespective of the cutoffs applied to the positive control cohort may be related to the mild nature of most cases included (17% asymptomatic, 57% mild, 9% moderate-severe, 13% critical based on WHO stratification criteria [18]). The relatively long time since PCR-confirmation (median: 181 days, range: 27-270) did not appear to be clearly related to antigen reactivity, in contrast to other reports [27]. Although use of such a positive control cohort adversely affects the apparent performance of the test, this combination of low-acuity illnesses and variable time since infection is likely to be reflective of a community serosurveillance sample and the sensitivity of the test was considered acceptable for population use. Further qualification in a larger group of confirmed Malian COVID-19 cases is planned.

While a threshold of three or four standard deviations above the mean of the negative control cohort is commonly used to establish specific preliminary cutoffs in single antigen assays, these cutoffs are prohibitively high in Mali and do not achieve a test performance appropriate for community serosurveillance. Furthermore, a threshold of two standard deviations above the mean of the negative control cohort, which was considered the optimal method in the US population [14], similarly reduced assay sensitivity to nearly 50% when using a two-antigen approach. In contrast, applying population-specific cutoffs derived from interrogation of ROC curves generated from local positive and negative control cohorts was able to improve test performance.

This methodical evaluation of serological assay options and the adaptation of approaches for use in Mali have yielded an optimized test that is well-validated and makes use of existing laboratory infrastructure. While increased background reactivity to SARS-CoV-2 antigens must be acknowledged, this reactivity may be largely offset through the use of a two-antigen assay and adaptation of assay cutoffs to suit the local population. Although the test characteristics remain imperfect, a thorough understanding of test performance will provide reassurance for estimates in future community serosurveillance.

## Data Availability

The data that support the findings of this study are available in the manuscript

## ACKNOWLEDGEMENTS

We gratefully acknowledge the support of Professor Yacouba Toloba (Point G Hospital) for assistance contacting convalescent cases for sample collection; Dr Mamady Kone (MRTC/USTTB), Emily Higbee and Jacquelyn Lane (LMIV/NIAID) for assistance coordinating the collection of convalescent samples; Dr. Thayne Dickey (LMIV/NIAID) for providing monoclonal neutralizing antibody CR3022; the Adventist Hospital, Maryland for providing US positive control convalescent samples used in the pseudovirus neutralization assay; Matthew Drew, Kelly Snead, Jennifer Mehalko, and Vanessa Wall (FNLCR) for production of antigens; Professor Boubacoar Traore (MRTC/USTTB) and Peter Crompton (MIBIU/NIAID) for providing samples from Kalifabougou; Patrick Gorres (LMIV/NIAID) for editorial assistance; the Malian COVID-19 Coordinator; and Ministry of Health for permission to partner in developing serosurveillance capacity in Mali.

## REFERENCES

1. Dyer, O., Covid-19: Many poor countries will see almost no vaccine next year, aid groups warn. BMJ, 2020. 371: p. m4809.

2. Bryant, J.E., et al., Serology for SARS-CoV-2: Apprehensions, opportunities, and the path forward. Sci Immunol, 2020. 5(47).

3. Theel, E.S., et al., Application, Verification, and Implementation of SARS-CoV-2 Serologic Assays with Emergency Use Authorization. J Clin Microbiol, 2020. 59(1).

4. Everett, D.B., et al., Association of schistosomiasis with false-positive HIV test results in an African adolescent population. J Clin Microbiol, 2010. 48(5): p. 1570–7.

5. Fonseca, M.O., et al., Cross-reactivity of anti-Plasmodium falciparum antibodies and HIV tests. Trans R Soc Trop Med Hyg, 2000. 94(2): p. 171–2.

6. Lejon, V., et al., Low specificities of HIV diagnostic tests caused by Trypanosoma brucei gambiense sleeping sickness. J Clin Microbiol, 2010. 48(8): p. 2836–9.

7. Tso, F.Y., et al., High prevalence of pre-existing serological cross-reactivity against severe acute respiratory syndrome coronavirus-2 (SARS-CoV-2) in sub-Saharan Africa. Int J Infect Dis, 2021. 102: p. 577–583.

8. Yadouleton, A., et al., Limited Specificity of Serologic Tests for SARS-CoV-2 Antibody Detection, Benin. Emerg Infect Dis, 2021. 27(1).

9. FDA. EUA Authorized Serology Test Performance. 2020 13 January 2021]; Available from: https://www.fda.gov/medical-devices/coronavirus-disease-2019-covid-19-emergency-use-authorizations-medical-devices/eua-authorized-serology-test-performance.

10. Esposito, D., et al., Optimizing high-yield production of SARS-CoV-2 soluble spike trimers for serology assays. Protein Expr Purif, 2020. 174: p. 105686.

11. Mehalko, J., et al., Improved production of SARS-CoV-2 spike receptor-binding domain (RBD) for serology assays. bioRxiv, 2020.

12. Hicks, J., et al., Serologic cross-reactivity of SARS-CoV-2 with endemic and seasonal Betacoronaviruses. medRxiv, 2020.

13. Aricescu, A.R., W. Lu, and E.Y. Jones, A time- and cost-efficient system for high-level protein production in mammalian cells. Acta Crystallogr D Biol Crystallogr, 2006. 62(Pt 10): p. 1243–50.

14. Klumpp-Thomas, C., et al., Standardization of ELISA protocols for serosurveys of the SARS-CoV-2 pandemic using clinical and at-home blood sampling. Nat Commun, 2021. 12(1): p. 113.

15. Wu, Y., et al., A noncompeting pair of human neutralizing antibodies block COVID-19 virus binding to its receptor ACE2. Science, 2020. 368(6496): p. 1274–1278.

16. Mukaka, M.M., Statistics corner: A guide to appropriate use of correlation coefficient in medical research. Malawi Med J, 2012. 24(3): p. 69–71.

17. Huang, A.T., et al., A systematic review of antibody mediated immunity to coronaviruses: kinetics, correlates of protection, and association with severity. Nat Commun, 2020. 11(1): p. 4704.

18. WHO, Clinical management of COVID-19. 27 May 2020.

19. Falzarano, D., et al., Dromedary camels in northern Mali have high seropositivity to MERS-CoV. One Health, 2017. 3: p. 41–43.

20. Mbow, M., et al., COVID-19 in Africa: Dampening the storm? Science, 2020. 369(6504): p. 624–626.

21. Njenga, M.K., et al., Why is There Low Morbidity and Mortality of COVID-19 in Africa? Am J Trop Med Hyg, 2020. 103(2): p. 564–569.

22. Premkumar, L., et al., The receptor binding domain of the viral spike protein is an immunodominant and highly specific target of antibodies in SARS-CoV-2 patients. Sci Immunol, 2020. 5(48).

23. Shi, R., et al., A human neutralizing antibody targets the receptor-binding site of SARS-CoV-2. Nature, 2020. 584(7819): p. 120–124.

24. Tehrani, Z.R., et al., Specificity and Performance of Nucleocapsid and Spike-based SARS-CoV-2 Serologic Assays. medRxiv, 2020.

25. McAndrews, K.M., et al., Heterogeneous antibodies against SARS-CoV-2 spike receptor binding domain and nucleocapsid with implications for COVID-19 immunity. JCI Insight, 2020. 5(18).

26. Yamaoka, Y., et al., Whole nucleocapsid protein of SARS-CoV-2 may cause false positive results in serological assays. Clin Infect Dis, 2020.

27. Dan, J.M., et al., Immunological memory to SARS-CoV-2 assessed for greater than six months after infection. bioRxiv 2020.

